# Mental health service activity during COVID-19 lockdown among individuals with Personality Disorders: South London and Maudsley data on services and mortality from January to May 2020

**DOI:** 10.1101/2020.09.13.20193730

**Authors:** Eleanor Nuzum, Evangelia Martin, Matthew Broadbent, Robert Stewart

## Abstract

The lockdown and social distancing policy imposed due to the COVID-19 pandemic is likely to have a widespread impact on mental healthcare for both services themselves and the people accessing those services. Previous reports from the South London and Maudsley NHS Trust (SLaM; a large mental health service provider for 1.2m residents in South London) highlighted a shift to virtual contacts among those accessing community mental health and home treatment teams and an increase in deaths over the pandemic’s first wave. However, there is a need to understand this further for specific groups, including those diagnosed with a personality disorder who might have particular vulnerabilities. Taking advantage of the Clinical Record Interactive Search (CRIS) data resource with 24-hourly updates of electronic mental health records data, this paper describes daily caseloads and contact numbers (face-to-face and virtual) for individuals with personality disorders across community, specialist, crisis and inpatient services. The report focussed on the period 1st January to 31st May 2020. We also report on daily accepted and discharged trust referrals, total trust caseloads and daily inpatient admissions and discharges for individuals with personality disorders. In addition, daily deaths are described for all current and previous SLaM service users with personality disorder over this period. In summary, comparing periods before and after 16th March 2020 there was a shift from face-to-face contacts to virtual contacts across all teams. Liaison and Older Adult teams showed the largest drop in caseloads, whereas Early Intervention in Psychosis service caseloads remained the same. Reduced accepted referrals and inpatient admissions were observed and there was a 28% increase in average daily deaths in the period after 16th March, compared to the period 1st January to 15th March.

## Background

The COVID-19 pandemic has had a significant impact across health services. Along with the impact of the virus itself, lockdown restrictions and social distancing policies may have had a direct impact on both psychological health and changes to service delivery. Services faced challenges in terms of increased staff sickness and self-isolation, suspected or confirmed COVID-19 infections in inpatient and outpatient settings, the need to minimise face-to-face contacts and the need to accommodate increasing pressures on acute medical care from cases of viral pneumonia. Changes in the availability and delivery of mental healthcare may have had a particular impact on individuals with pre-existing mental health conditions. Within this group, individuals diagnosed with a personality disorder tend to access support from a range of services, including inpatient, community and crisis services all of which have rapidly reconfigured the way they operate following lockdown. The impacts of social isolation for this patient group are as yet unclear, along with the impact of changes to services accessed, and it is important to increase information in the public domain (1).

We have previously begun reporting on the mental healthcare impact of the UK COVID-19 pandemic, taking advantage of the Clinical Record Interactive Search (CRIS) data platform that receives 24-hourly updates from its source electronic mental health records. Previous pandemic era reports are listed on https://www.maudsleybrc.nihr.ac.uk/facilities/clinical-record-interactive-search-cris/covid-19-publications/. By looking at activity in services January-May 2020, it is possible to compare activity before and after 16th March, the date from which lockdown was imposed by the government. We have previously reported an overall shift from face-to-face to virtual contacts in community mental health teams and home treatment teams, as well as a drop in caseloads and total contacts in home treatment teams (2). We are seeking to extend this analysis to groups who might have specific needs and service use characteristics that might not be well reflected in Trust-wide data, and this report focuses on service users with a personality disorder diagnosis.

## Methods

The Biomedical Research Centre (BRC) Case Register at the South London and Maudsley NHS Foundation Trust (SLaM) has been described previously (3;4). SLaM serves a geographic catchment of four south London boroughs (Croydon, Lambeth, Lewisham, Southwark) with a population of around 1.2 million residents, and has used a fully electronic health record (EHR) across all its services since 2006. SLaM’s BRC Case Register was set up in 2008, providing researcher access to de-identified data from SLaM’s EHR via the Clinical Record Interactive Search (CRIS) platform and within a robust security model and governance framework (5). CRIS has been extensively developed over the last 10 years with a range of external data linkages and natural language processing resources (6). CRIS is updated from SLaM’s EHR every 24 hours and thus provides relatively ‘real-time’ data. SLaM’s EHR is itself immediately updated every time an entry is made, which include date-stamped fields indicating patient contacts (‘events’) and those indicating acceptance of a referral or a discharge from a given service (or SLaM care more generally). Mortality in the complete EHR (i.e. all SLaM patients with records, past or present) is ascertained weekly through automated checks of National Health Service (NHS) numbers (a unique identifier used in all UK health services) against a national spine. CRIS has supported over 200 peer reviewed publications to date. CRIS has received approval as a data source for secondary analyses (Oxford Research Ethics Committee C, reference 18/SC/0372).

Activity and caseload data were extracted via CRIS and enumerated for every day from 1st January 2020 to 31st May 2020. The focus of this report is on clinical activity and mortality of individuals with a previous diagnosis of a personality disorder; this was defined as ICD-10 codes F60x or F61x recorded within structured fields for primary or secondary diagnoses in the source electronic health record (mandatory fields for all active caseloads), supplemented by ‘personality disorder’ ascertained from a bespoke natural language processing algorithm developed within CRIS to extract text associated with diagnostic statements recorded in open text fields (https://www.maudsleybrc.nihr.ac.uk/facilities/clinical-record-interactive-search-cris/cris-natural-language-processing/). This approach allowed for the fact that personality disorder is under-reported in coded diagnostic fields in favour of other co-occurring diagnoses.

Clinical activity was measured across 6 service areas; working age community Adult Mental Health Services (AMHS), Child and Adolescent Mental Health Services (CAMHS), Mental Health of Older Adults (MHOA), Early Intervention in Psychosis (EIP), Home Treatment Team and crisis services (HTT), and Liaison services including other services covering Accident & Emergency referrals (Liaison/A&E). Total Trust caseloads, number of accepted and discharged Trust referrals, number of inpatient admissions and discharges, and number of inpatients (total and those specifically under a Mental Health Act order) were also extracted for the client group.

For AMHS, CAMHS, MHOA, EIP, HTT, Liaison, daily caseloads were calculated by ascertaining patients who were receiving active care from the service on a given day, based on the date a referral to that service was recorded as accepted to the point a discharge was made from that service. Daily contact numbers were ascertained from recorded ‘events’ (i.e. standard case note entries) for that service and were divided into the following groups according to structured compulsory meta-data fields for that event in the EHR: i) face-to-face contacts attended; ii) virtual contacts attended (by email, fax, mail, phone, online, or video link); iii) total contacts, as the sum of these two; iv) did not attend (DNA). Finally, mortality data (number of deaths for all patients with personality disorder with SLaM records) were extracted for the period in question. Data extractions were carried out on 5th June 2020.

Descriptive data were generated daily for the above parameters and displayed graphically. Weekend days and national holidays were omitted for AMHS, CAMHS, MHOA, and EIP contacts. For comparisons between time periods, lockdown was defined as commencing on 16th March 2020, the date this policy was announced by the UK government. Mean (SD) activity levels were described before and after this date and percentage changes quantified.

## Results

Mean daily statistics before and after 16th March and proportional changes are displayed in Table 1. Daily deaths in the period from 1st January to 31st May are displayed in Figure 1 and weekly deaths in Figure 2. Mean daily deaths increased by 28% in the period after 16th March compared to the period between 1st January to 15th March.

**Table 1.** Levels and changes in daily service caseloads and contacts in patients with a personality disorder diagnosis from 1st January to 31st May 2020

| Service metric | Mean (SD) daily level before<br>16<sup>th</sup> March | Mean (SD) daily level from<br>16<sup>th</sup> March | Difference | Percentage change |
| --- | --- | --- | --- | --- |
| Deaths | 0.56 (0.75) | 0.72 (0.90) | 0.16 | +28.1% |
| Total Trust caseload | 4823.1 (13.5) | 4644.0 (89.4) | -179.1 | -3.7% |
| Accepted Trust referrals | 15.8 (4.8) | 10.0 (3.7) | -5.7 | -36.3% |
| Discharged from Trust services | 16 (3.1) | 15.4 (5.1) | -0.6 | -4.0% |
| Inpatient admissions | 3.7 (1.8) | 2.1 (1.1) | -1.6 | -42.6% |
| Discharges from inpatient care | 4.4 (2.3) | 3.8 (2.4) | -0.6 | -13.3% |
| Inpatient numbers | 238.8 (5.6) | 183.6 (15.4) | -55.1 | -23.1% |
| Inpatient numbers under MHA | 185.3 (5.9) | 150.5 (10.6) | -34.8 | -18.8% |
| AMHS total contacts | 185.4 (21) | 169 (21.4) | -16.4 | -8.9% |
| AMHS face-to-face contacts | 136.4 (20.1) | 45.0 (14.6) | -91.4 | -67% |
| AMHS virtual contacts | 49 (7.4) | 124 (20.4) | 75 | +152.9% |
| AMHS caseload | 2170.9 (10.6) | 2126.3 (27.9) | -44.7 | -2.1% |
| CAMHS total contacts | 16.4 (5.1) | 14.4 (3.6) | -2.0 | -12.3% |
| CAMHS face-to-face contacts | 10.7 (3.5) | 1.5 (1.6) | -9.2 | -85.9% |
| CAMHS virtual contacts | 5.7 (2.9) | 12.9 (3.2) | 7.2 | +126.0% |
| CAMHS caseload | 216 (4.3) | 197.0 (9.8) | -19.0 | -8.8% |
| MHOA total contacts | 9.3 (3.1) | 6.9 (2.9) | -2.4 | -25.8% |
| MHOA face-to-face contacts | 6.5 (2.6) | 2.2 (1.7) | -4.3 | -66.7% |
| MHOA virtual contacts | 2.8 (1.5) | 4.7 (2.3) | 1.9 | +67.9% |
| MHOA caseload | 94.4 (4.2) | 82.3 (5.6) | -12.1 | -12.8% |
| EIP total contacts | 8.9 (3.3) | 8.9 (3.4) | 0.0 | 0.0% |
| EIP face-to-face contacts | 5.7 (2.9) | 1.1 (1.5) | -4.6 | -81.0% |
| EIP virtual contacts | 3.3 (2.0) | 7.9 (2.8) | 4.6 | +140.5% |
| EIP caseload | 60.6 (0.8) | 58.3 (1.0) | -2.4 | -3.9% |
| HTT total contacts | 45.1 (7.4) | 26.0 (5.7) | -19.1 | -42.4% |
| HTT face-to-face contacts | 33.7 (5.8) | 11.1 (4.6) | -22.6 | -67.1% |
| HTT virtual contacts | 8.2 (3.6) | 12.5 (4.3) | 4.3 | +52.2% |
| HTT caseload | 55.4 (3.9) | 31.2 (9.1) | -24.2 | -43.6% |
| Liaison/A&E total contacts | 27.6 (6.0) | 19.4 (5.9) | -8.2 | -29.8% |
| Liaison/A&E face-to-face contacts | 25.3 (6.0) | 16.3 (5.4) | -9.1 | -35.7% |
| Liaison/A&E virtual contacts | 2.3 (1.8) | 3.1 (1.6) | 0.8 | +35.9% |
| Liaison/A&E caseload | 78.4 (4.4) | 63.0 (4.7) | -15.4 | -19.6% |

Abbreviations: AMHS working age adult mental health services, CAMHS child and adolescent mental health service, MHOA mental health of older adults, EIP early intervention for psychosis, HTT home treatment team.

**Figure 1.**
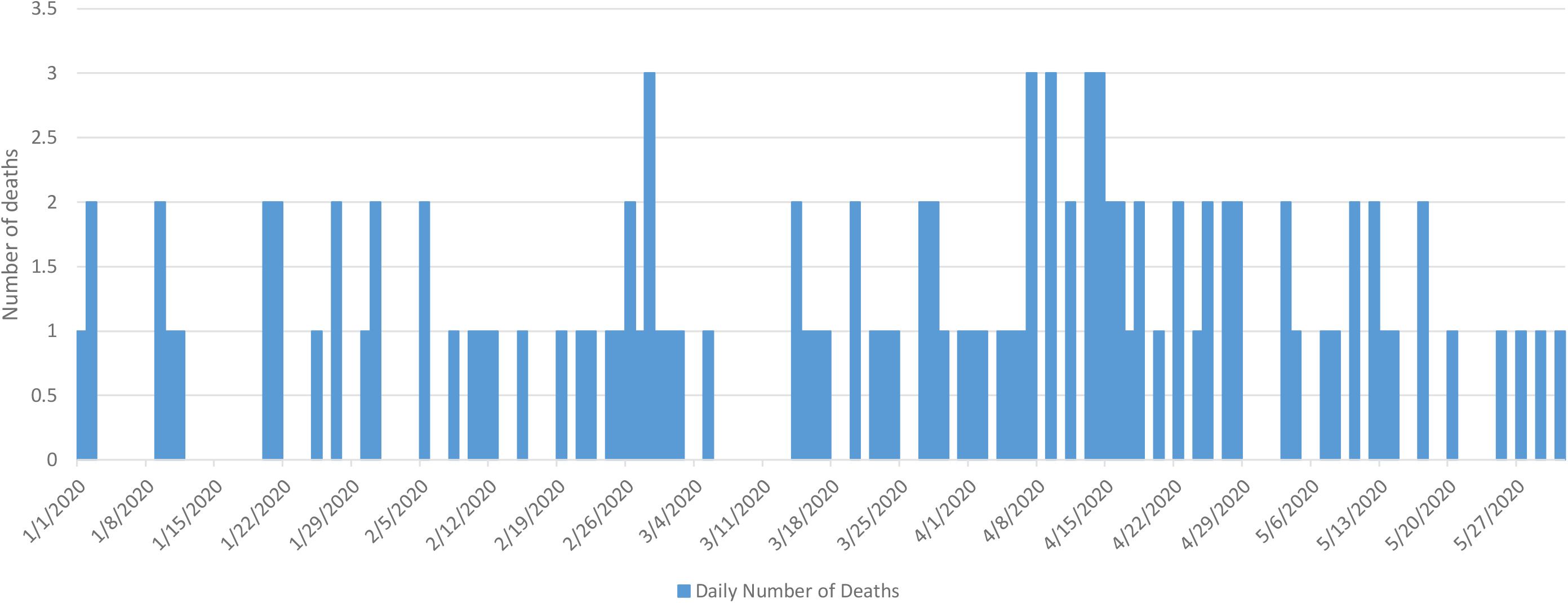
Daily recorded deaths for individuals diagnosed with personality disorder (daily; January - May 2020)

**Figure 2.**
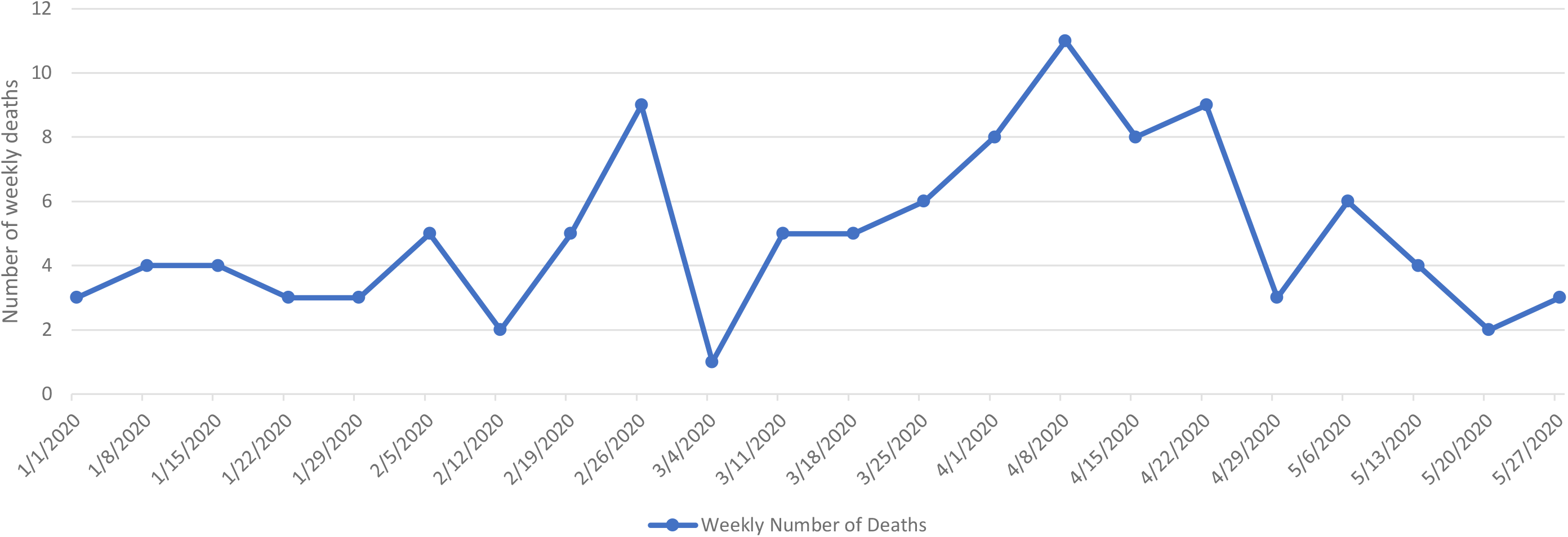
Recorded deaths for individuals diagnosed with personality disorder (weekly; January - May 2020)

The Trust total caseload for service users with personality disorders is displayed in Figure 3, and daily accepted and discharged trust referrals are displayed in Figure 4. The latter showed higher numbers of discharges compared to admissions from mid-March to late-April; however, overall numbers of admissions and discharges were very small compared to the total caseload numbers, which did not change substantially (4% lower after 15th March compared to beforehand).

**Figure 3.**
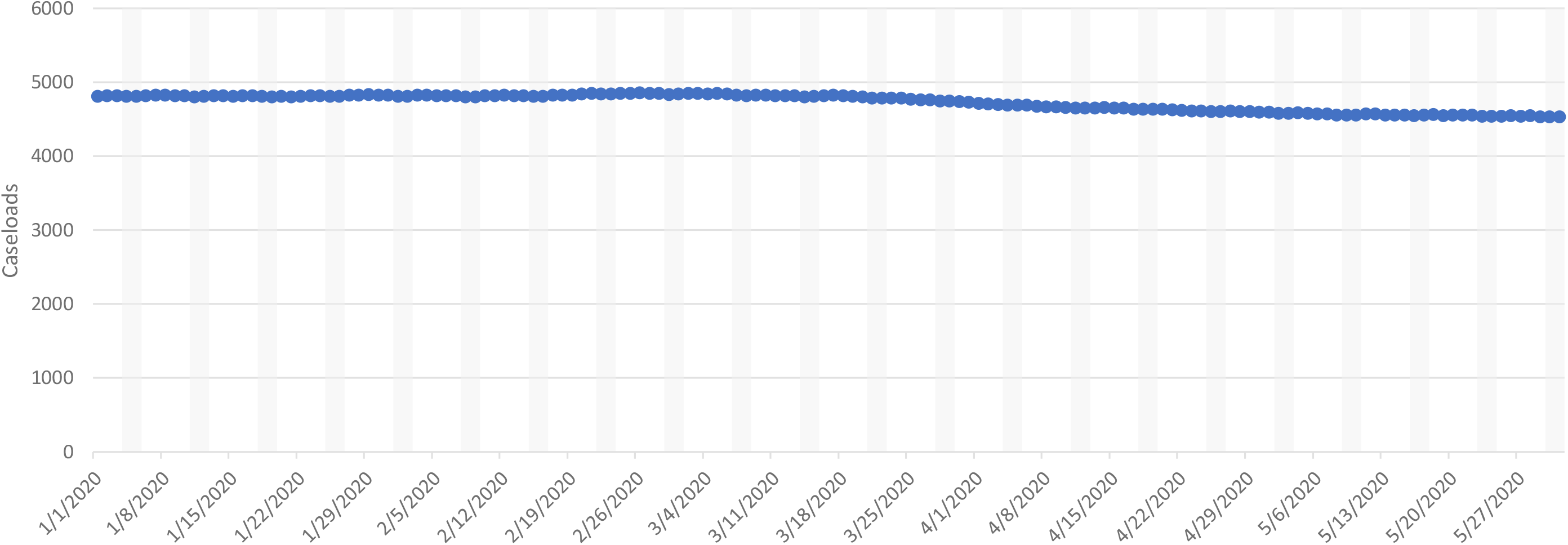
Trust caseloads of patients with a pesonality disorder diagnosis (daily; January - May 2020)

**Figure 4.**
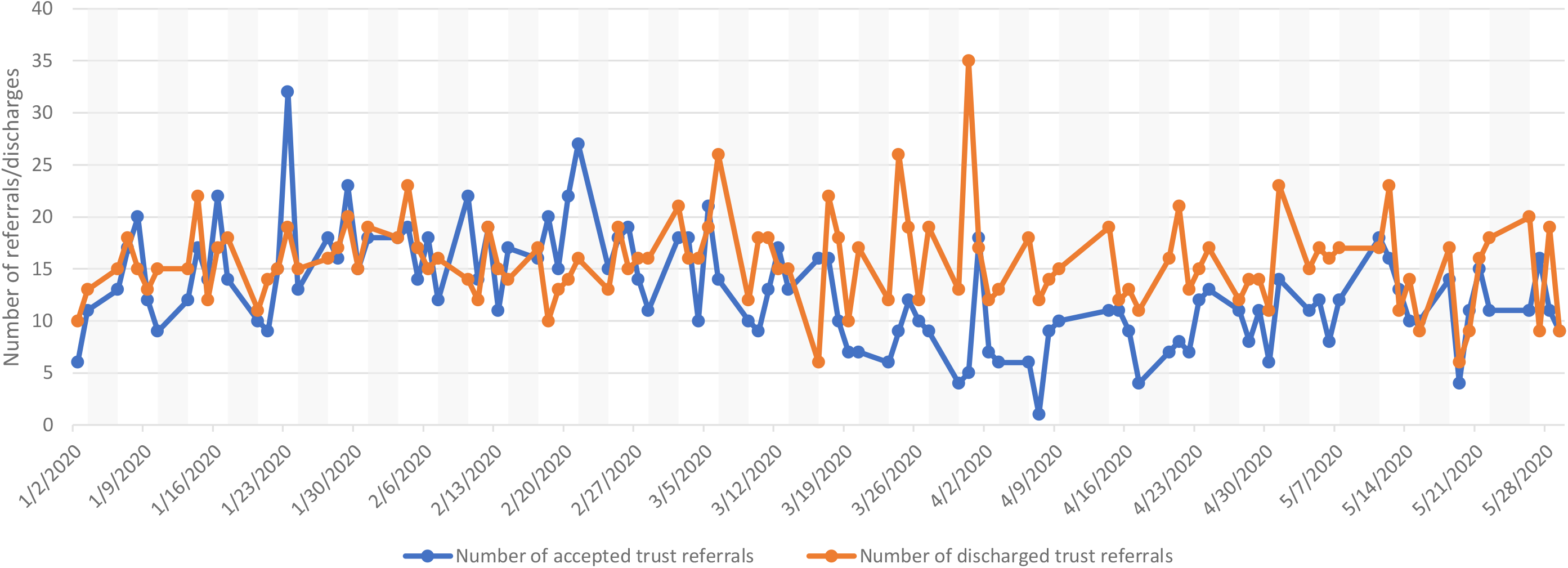
Number of accepted and discharged Trust referrals of patients with a pesonality disorder diagnosis (daily, January - May 2020)

Data on inpatient care are displayed in Figures 5-6, showing a fall in daily inpatient numbers (by 23% overall, by 19% for those under the Mental Health Act), accounted for by a greater decrease in admissions (43%) compared to discharges (13%) since 15th March.

**Figure 5.**
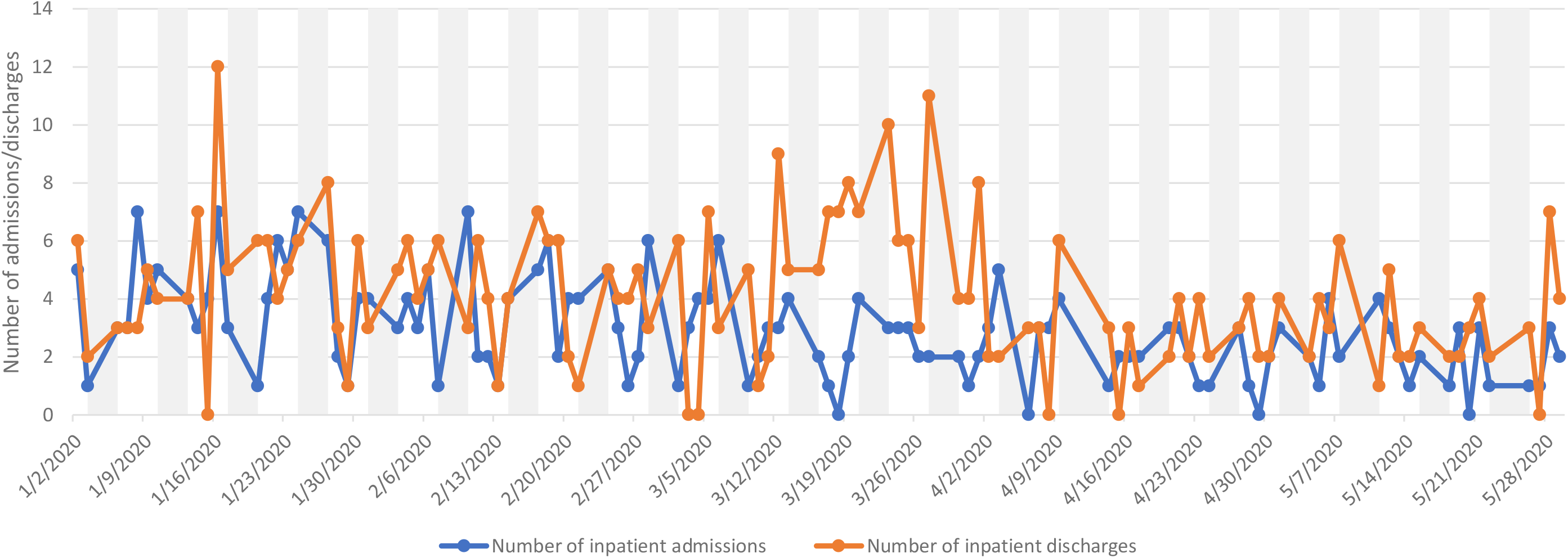
Inpatient admissions and discharges of patients with a pesonality disorder diagnosis (daily, January - May 2020)

**Figure 6.**
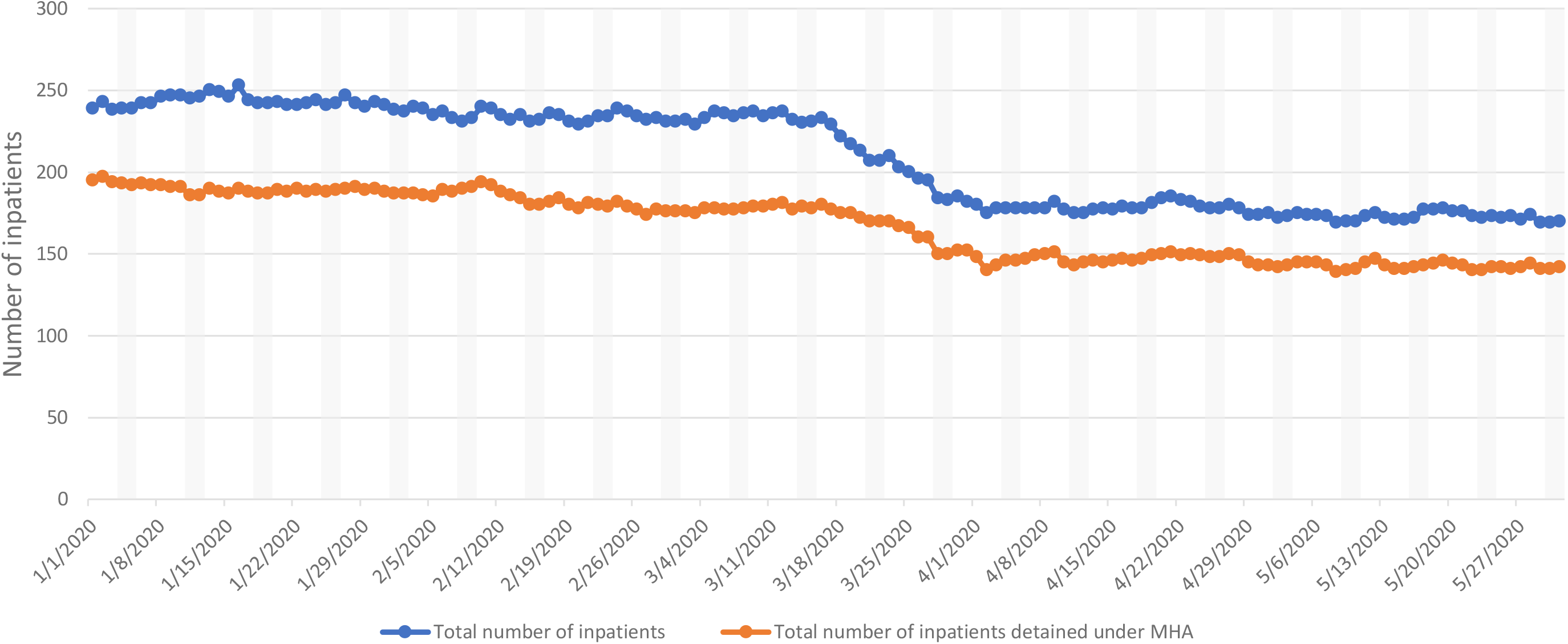
Total number of inpatients with a personality disorder diagnosis (daily; January-May 2020)

AMHS daily contacts are displayed in Figure 7 and daily caseloads are displayed in Figure 8. Mean total daily contacts showed a 9% reduction overall after 15th March, although levels by late May appear similar to those in January and February. Face-to-face contacts were reduced by 67% reduction, and virtual contacts were increased by 153%. Mean daily caseloads were relatively unchanged (2% reduced).

**Figure 7.**
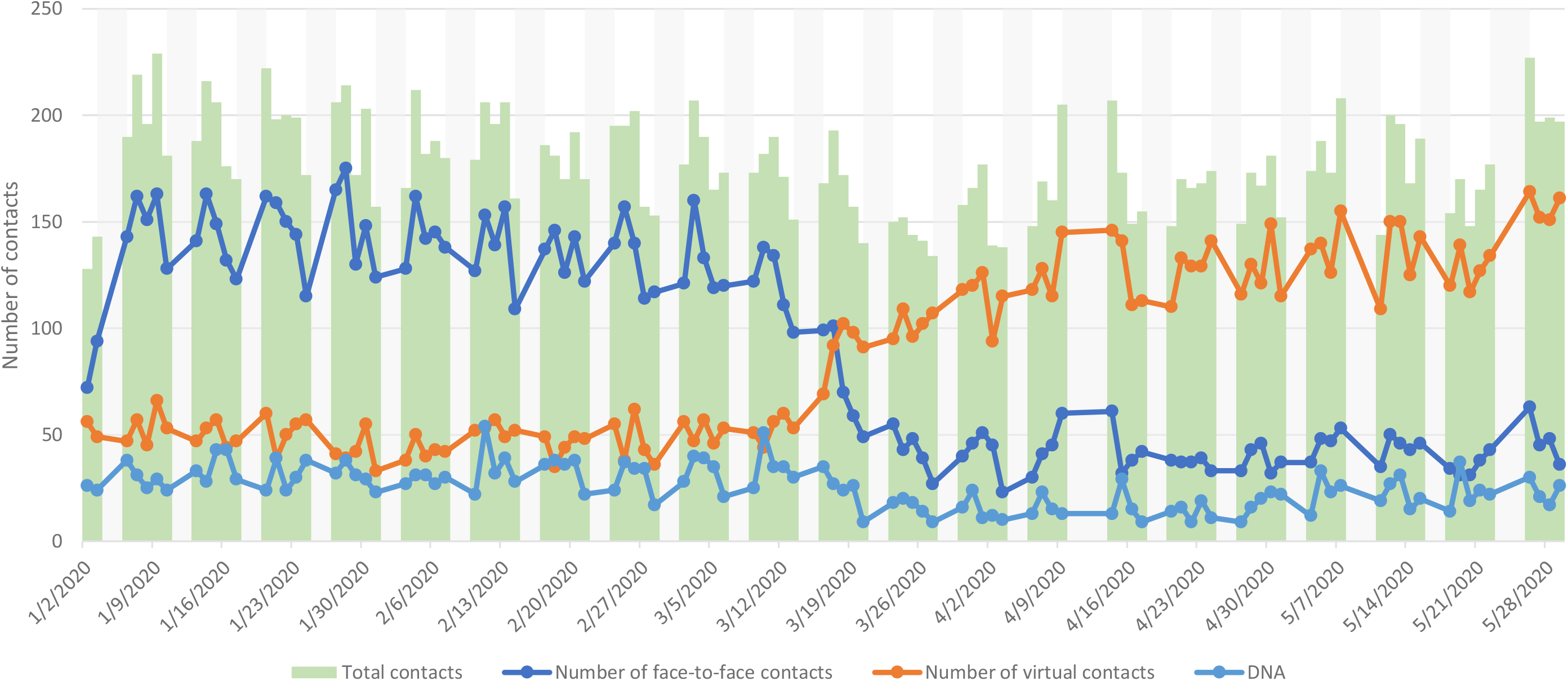
Working age adult psychiatry: non-specialist community service contacts by type (daily; January - Mary 2020) in patients with a pesonality disorder diagnosis

**Figure 8.**
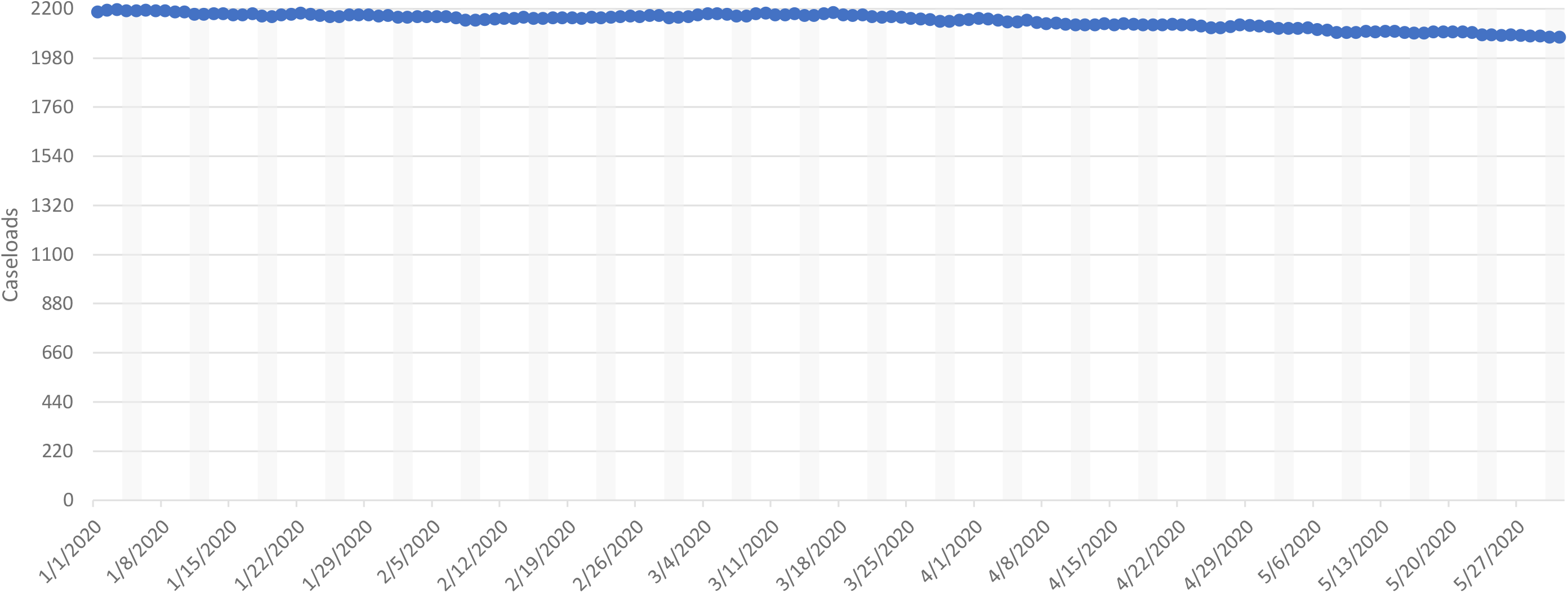
Working age adult psychiatry: non-specialist community services caseloads (daily; January - Mary 2020) of patients with a pesonality disorder diagnosis

CAMHS daily contacts are displayed in Figure 9 and daily caseloads are displayed in Figure 10. Mean total daily contacts were reduced by 12% overall after 15th March, made up of an 86% reduction in face-to-face contacts and a 126% increase in virtual contacts. Mean daily caseloads were reduced by 9%.

**Figure 9.**
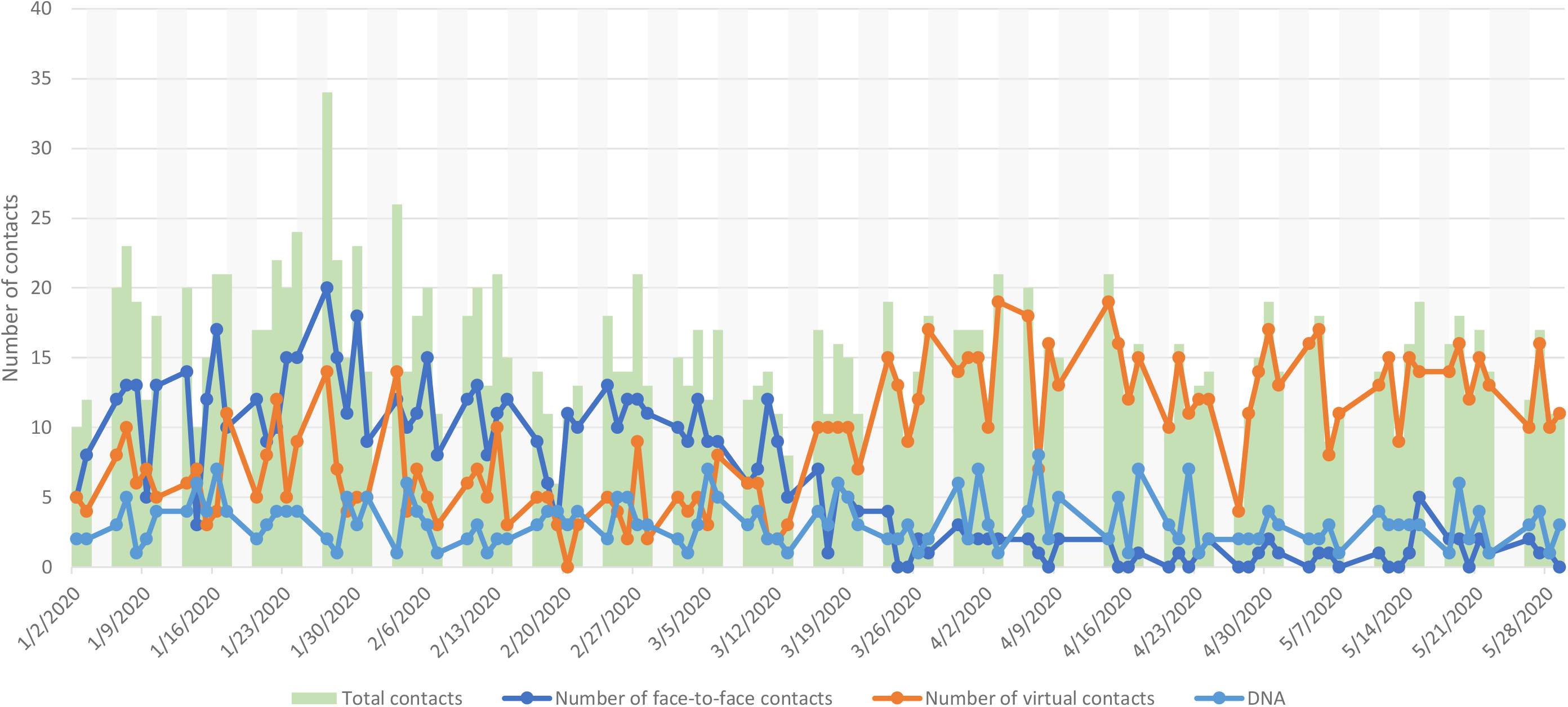
Child and Adolescent Mental Health Services contacts by type (daily; January - May 2020) for patients with a pesonality disorder diagnosis

**Figure 10.**
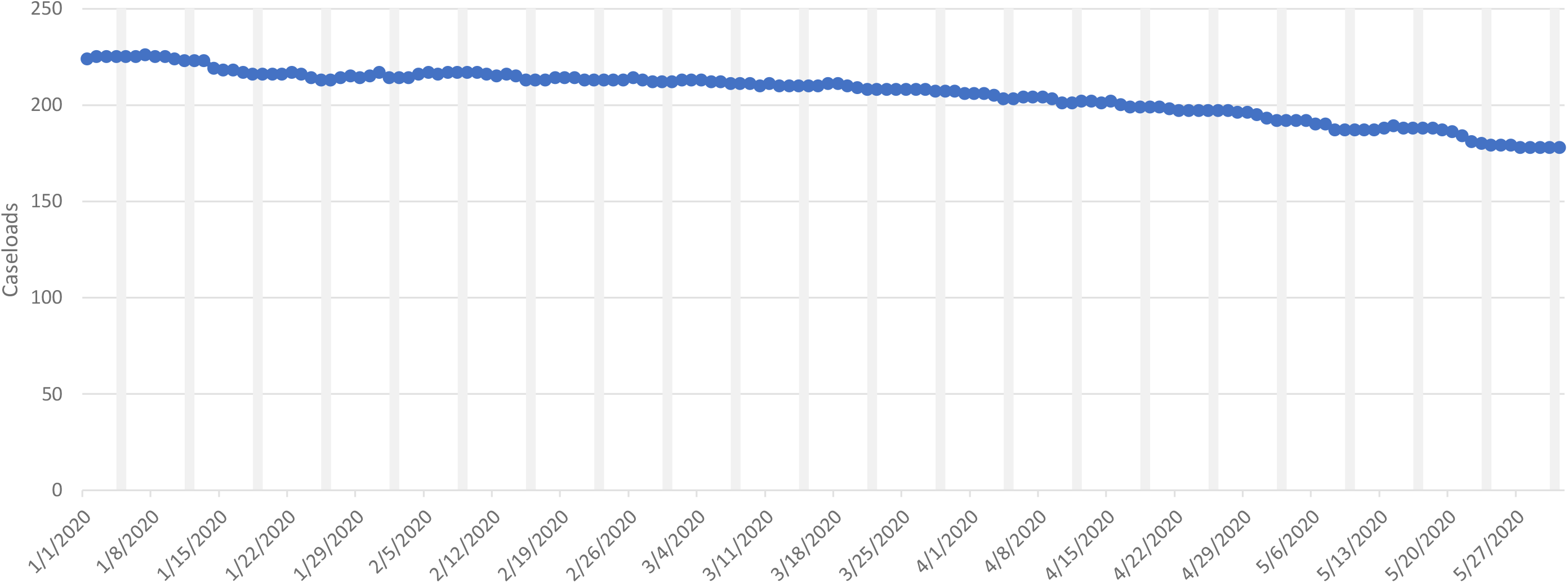
Child and Adolescent Mental Health Services caseloads (daily; January - May 2020) for patients with a pesonality disorder diagnosis

MHOA daily contacts are displayed in Figure 11 and daily caseloads are displayed in Figure 12. Mean total daily contacts were reduced by 26%: a 66.7% reduction in face-to-face contacts, and a 68% increase in virtual contacts. Mean daily caseloads were reduced by 13%.

EIP daily contacts are displayed in Figure 13 and daily caseloads are displayed in Figure 14. Mean total daily contacts showed no overall change: an 81% reduction in face-to-face contacts balanced by a 141% increase in virtual contacts. Mean daily caseloads showed a 4% reduction.

**Figure 11.**
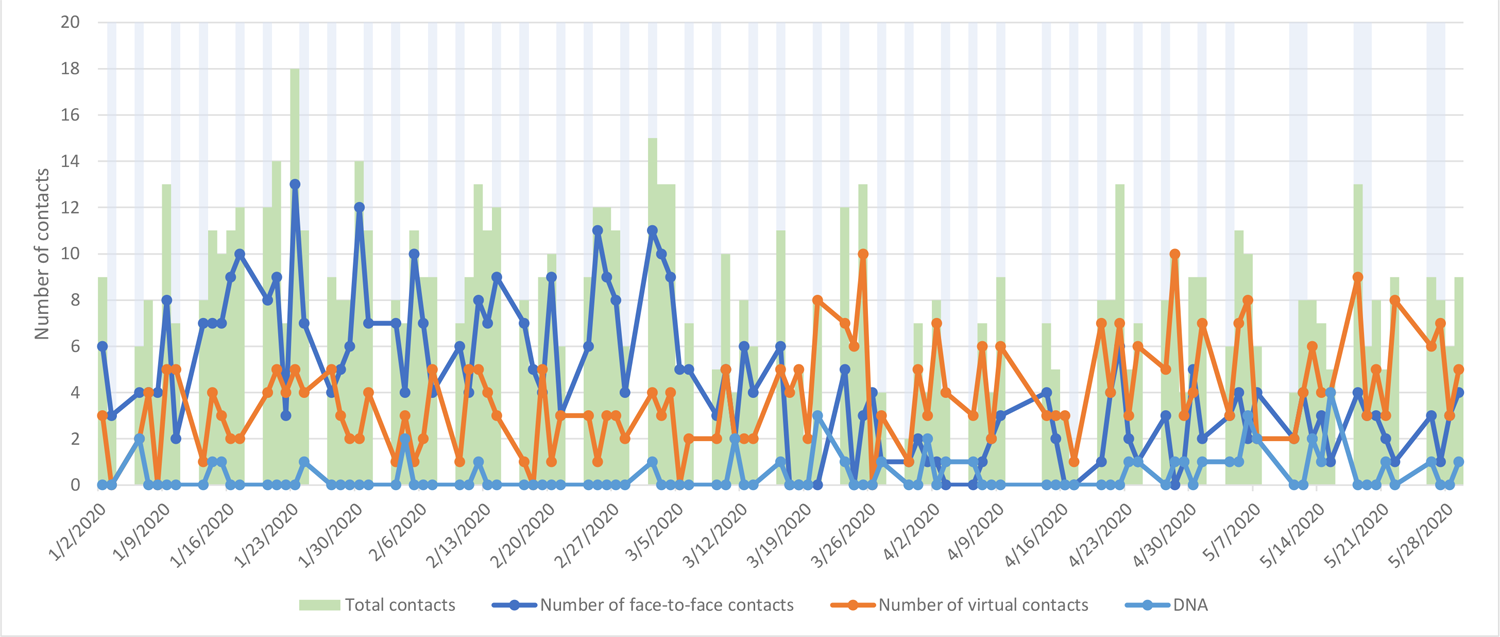
Older adults services contact by type (daily; January - May 2020) for patients with a pesonality disorder diagnosis

**Figure 12.**
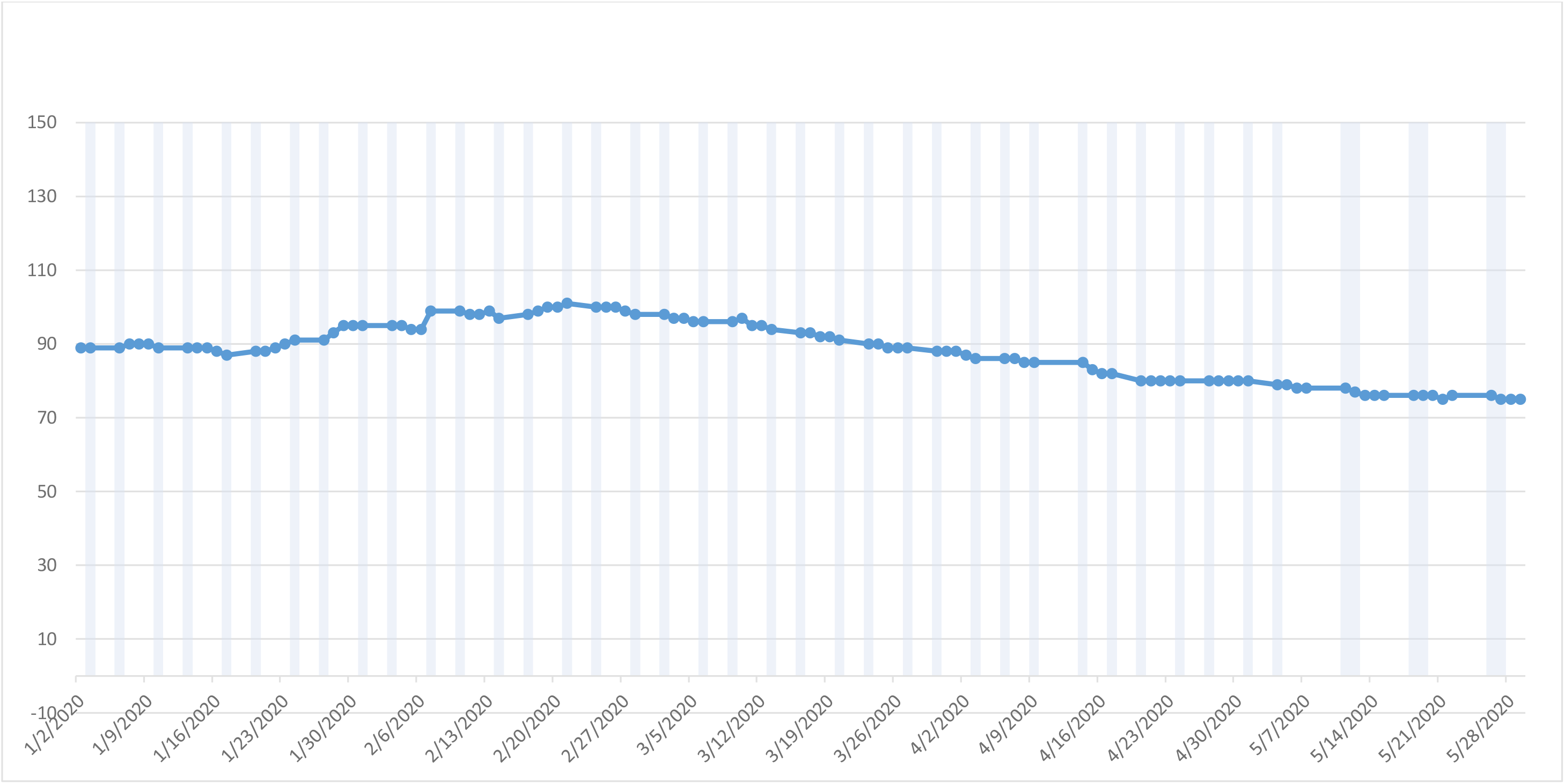
Older adult services caseloads (daily; January - May 2020) for patients with a pesonality disorder diagnosis

**Figure 13.**
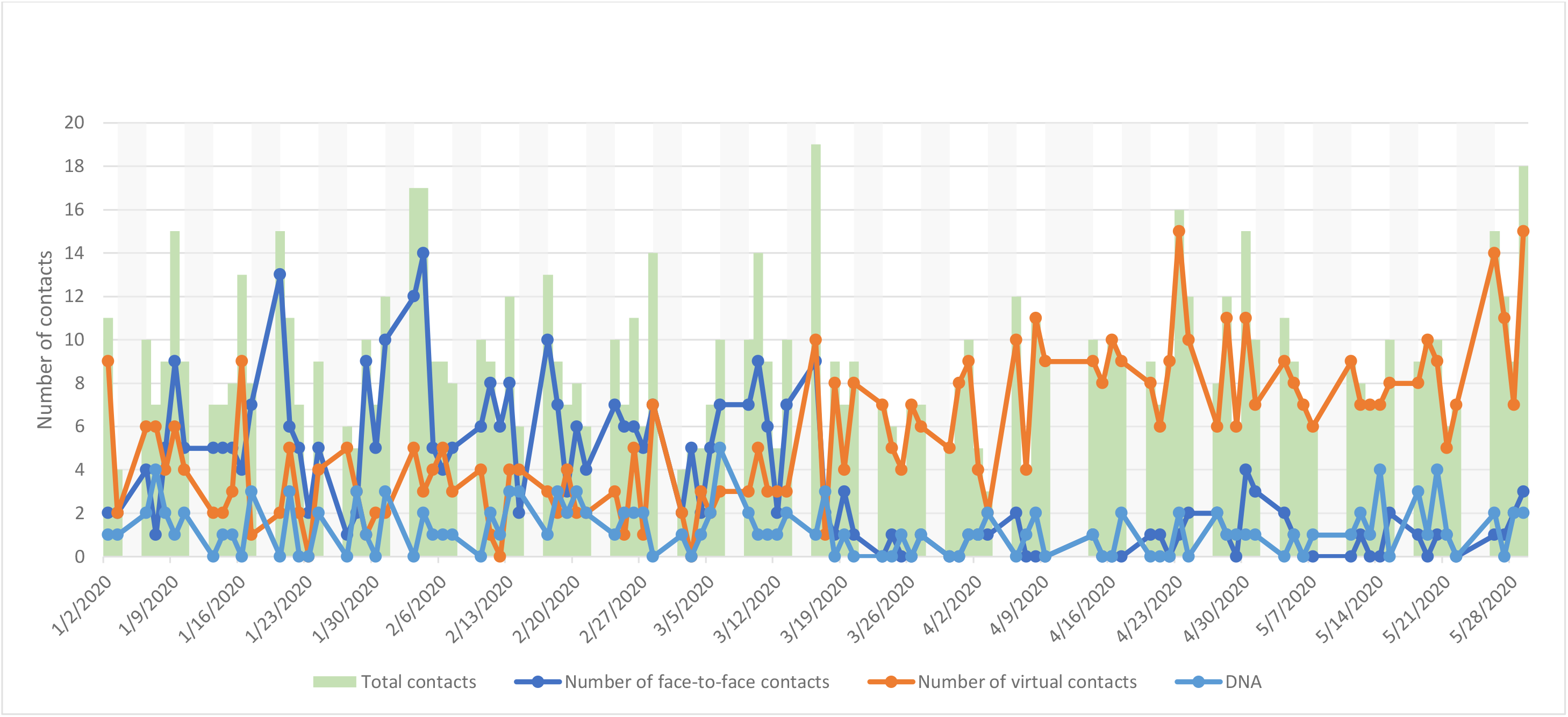
Early intervention psychosis service contacts by type (daily; January - May 2020) for patients with a pesonality disorder diagnosis

**Figure 14.**
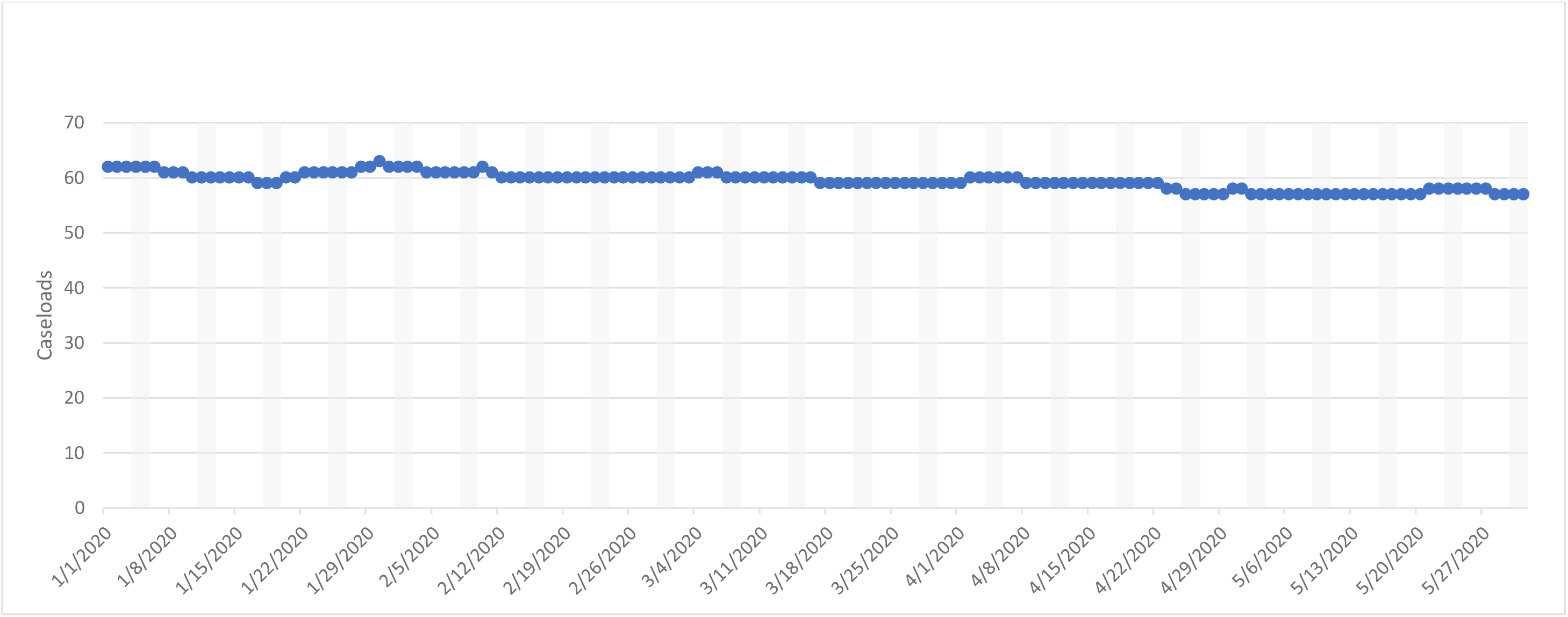
Early intervention psychosis service caseloads (daily; January - May 2020) for patients with a pesonality disorder diagnosis

HTT daily contacts are displayed in Figure 15 and daily caseloads are displayed in Figure 16. Mean total daily contacts were reduced by 42%: a 67% reduction in face-to-face contacts and a 53% increase in virtual contacts. Mean daily caseloads reduced by 44%.

**Figure 15.**
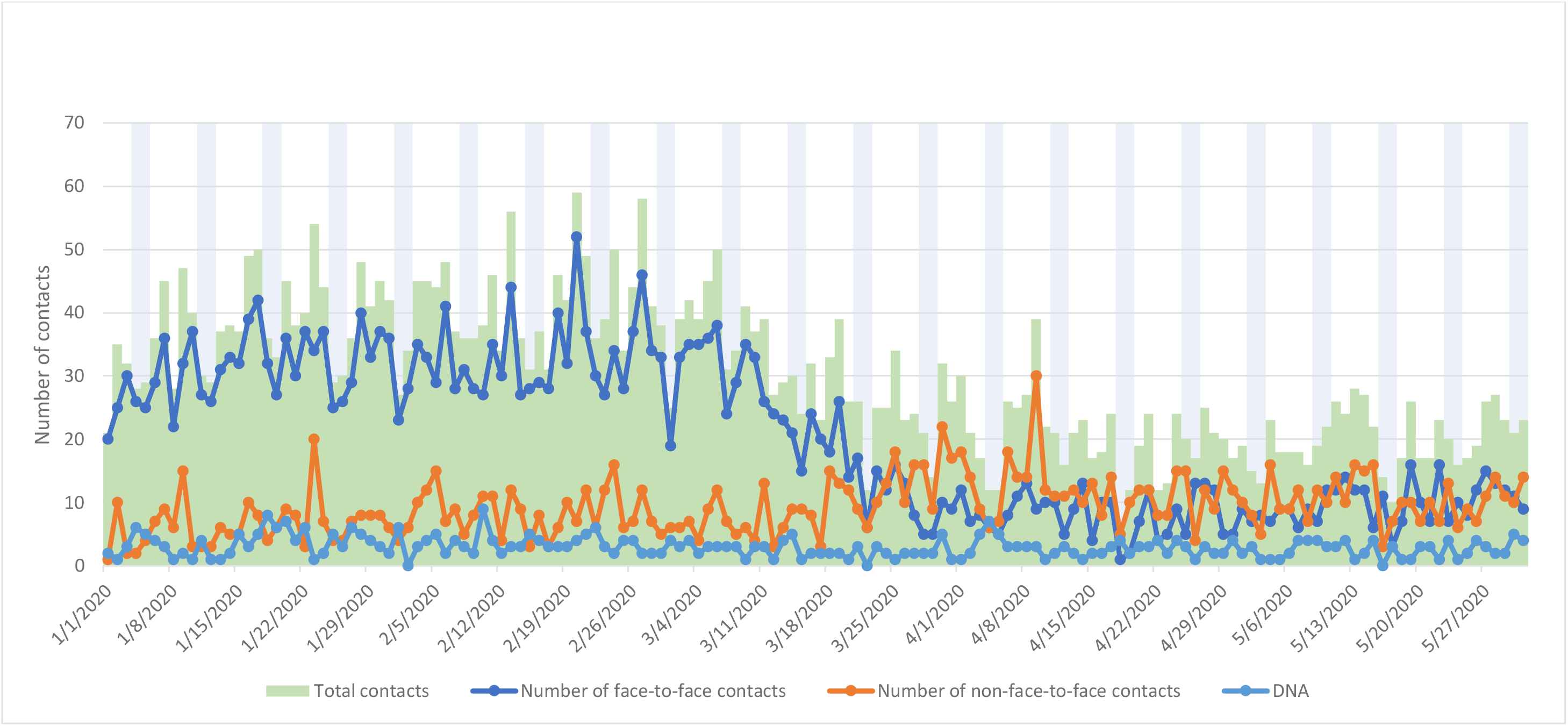
Home treatment / crisis service contacts by type (daily; January - May 2020) for patients with a pesonality disorder diagnosis

**Figure 16.**
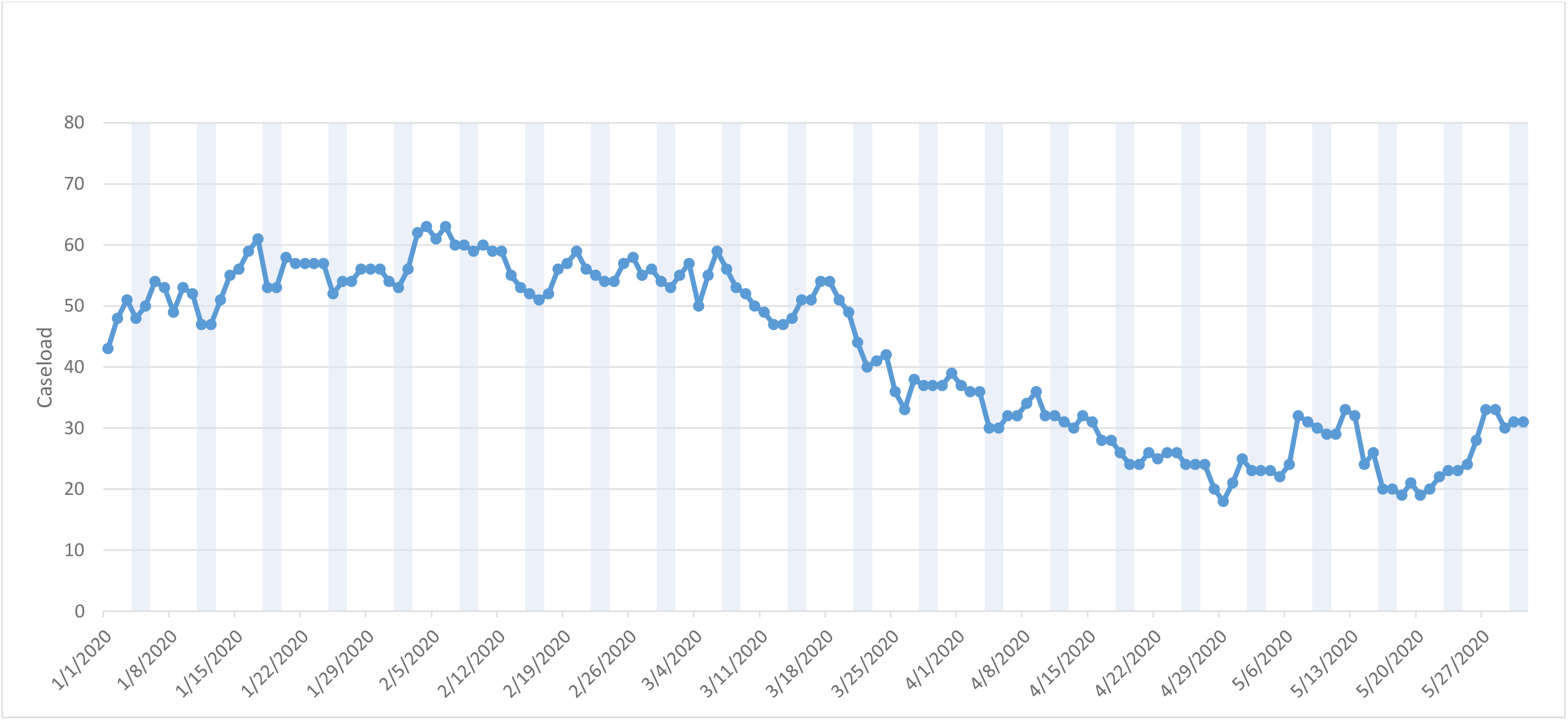
Home treatment / crisis services caseloads (daily; January - May 2020) for patients with a pesonality disorder diagnosis

Liaison/A&E daily contacts are displayed in Figure 17 and daily caseloads are displayed in Figure 18. Mean total daily contacts were reduced by 30%: a 35.7% reduction in face-to-face contacts and a 36% increase in virtual contacts. Mean daily caseloads were reduced by 20%.

**Figure 17.**
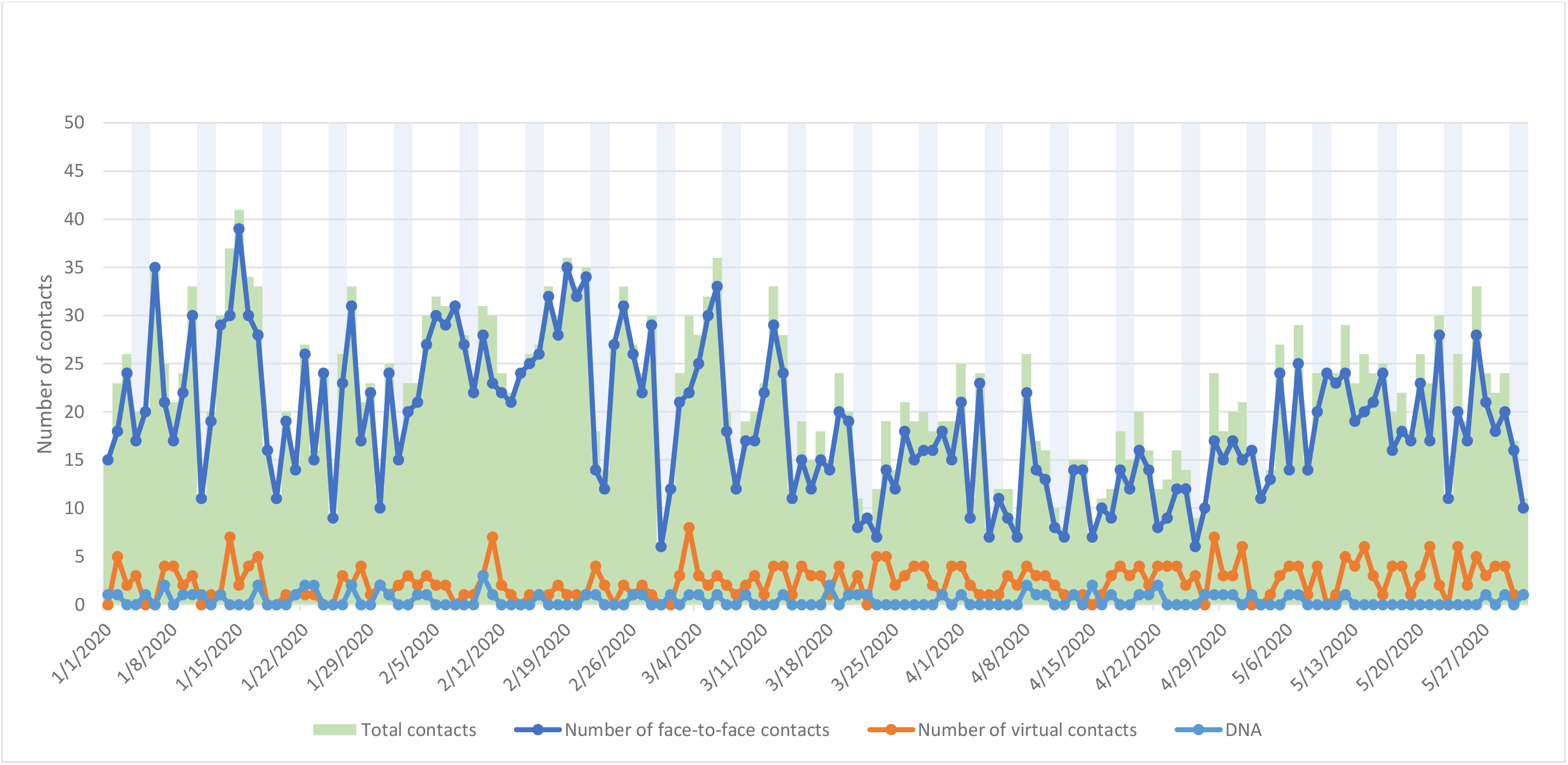
Liasion / A&E services contacts by type (January - May 2020) for patients with a pesonality disorder diagnosis

**Figure 18.**
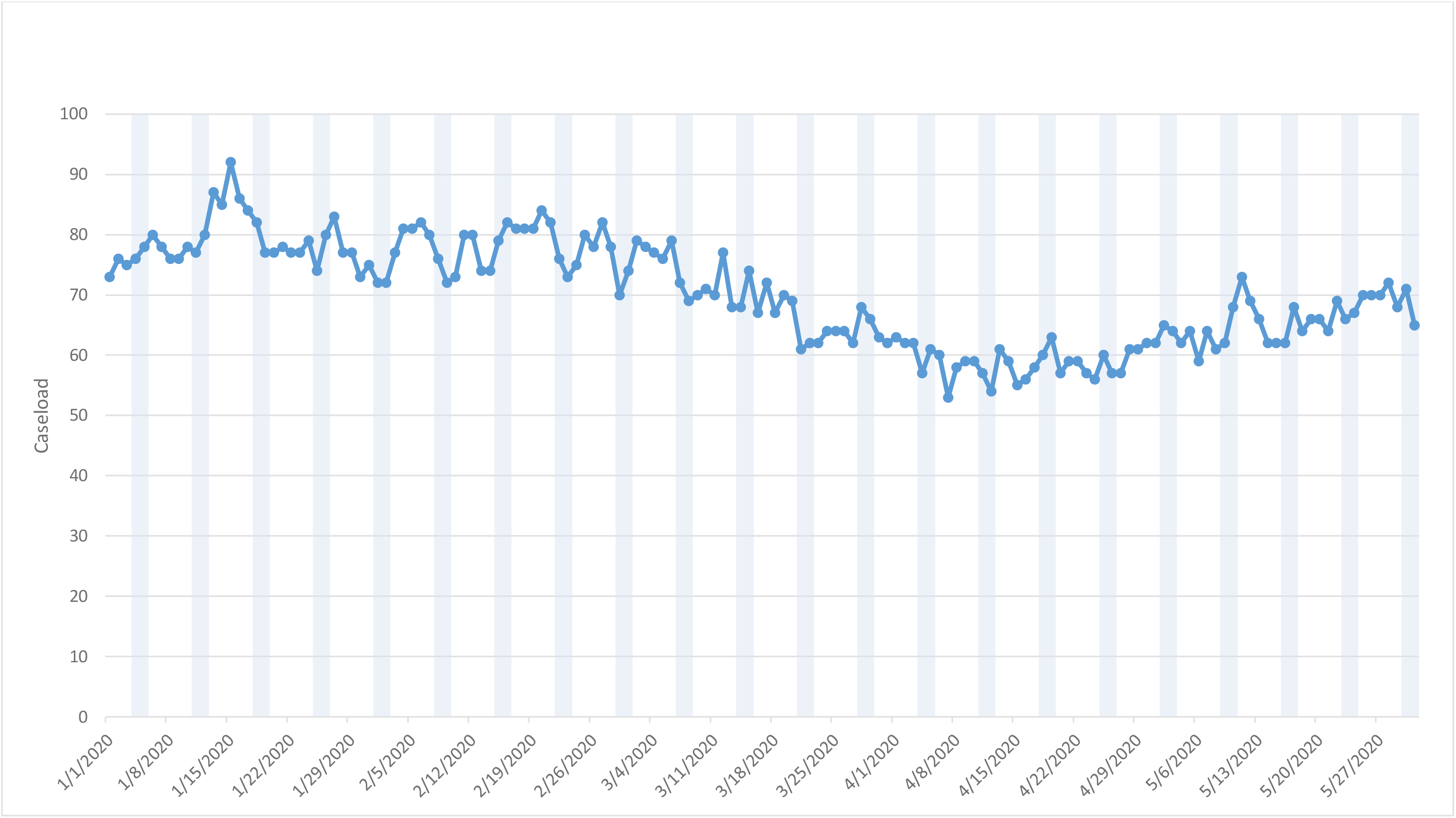
Liasion / A&E services caseloads (daily; January - May 2020) for patients with a pesonality disorder diagnosis

## Discussion

Considering patients with a previous personality disorder diagnosis receiving care from the South London and Maudsley Trust, comparing periods before and after the 16th March 2020 as the commencement of lockdown policy, daily caseloads showed slight reductions for AMHS, CAMHS and EIP (2%, 9% and 4% respectively), with larger reductions seen for MHOA, HTT and Liaison/A&E services (13%, 44% and 20% respectively). Total contacts did not change for EIP services, and there was a small decrease for AMHS (9%), while more noticeable reductions in total contacts were seen for CAMHS, MHOA, HTT and Liaison/A&E service groups (12%, 26%, 42% and 30% respectively). The largest changes observed were the decreases in face-to-face contacts and increases in virtual contacts across all services, with AMHS (face-to-face decrease by 67%, virtual contact increase by 153%), CAMHS (face-to-face decrease by 86%, virtual contact increase by 126%) and EIP (face-to-face decrease by 81%, virtual contact increase by 141%) services showing the most noticeable changes. Mean daily deaths for the period after 16th March increased by 28% compared to the period from 1st January to 15th March.

Considering the Trust as a whole, there was a large decrease in the number of accepted referrals (by 36%) and a small decrease in the number of discharged patients (by 4%), but these numbers were relatively small compared to the total caseload which was relatively unchanged (4% decrease). A marked decrease in inpatient admissions (by 43%) and a smaller decrease in inpatient discharges (by 14%) accompanied a decrease by 23% in the total number of inpatients in this diagnostic group after 16th March (18% in the number of inpatients detained under the Mental Health Act).

Considering limitations, it is important to bear in mind that the data are derived from a single site. Because complete data are being provided for that site with no hypothetical source population intended, calculation of confidence intervals was not felt to be appropriate for the descriptive data provided in this report; applicability to other mental healthcare providers cannot therefore be inferred and would need specific investigation. Profiles of services used by individuals with personality disorders and catchment morbidity are also likely to vary. It is also likely that we did not capture all of the services individuals with personality disorders are currently accessing for mental healthcare, as well as undiagnosed individuals. The diagnostic group defined for this study importantly refers to any previous diagnosis of a personality disorder and not necessarily the most recent diagnosis. Classification of service types is inevitably approximate and likely to vary between mental health service providers.

## Data Availability

The source data extract is available on request, subject to approval of the data owner (South London and Maudsley NHS Foundation Trust).

